# Estimating the optimal age for infant measles vaccination

**DOI:** 10.1101/2023.11.20.23298759

**Authors:** Elizabeth Goult, Laura Andrea Barrero Guevara, Michael Briga, Matthieu Domenech de Cellès

## Abstract

The persistence of measles in many regions demonstrates large immunity gaps, resulting from incomplete or ineffective immunization with measles-containing vaccines (MCVs). A key factor affecting MCV impact is age, with infants receiving dose 1 (MCV1) at older ages having a reduced risk of vaccine failure, but also an increased risk of contracting infection before vaccination. Here, we designed a new method—based on a transmission model incorporating realistic vaccination delays and age variations in MCV1 effectiveness—to capture this risk trade-off and estimate the optimal age for recommending MCV1. We predict a large heterogeneity in the optimal ages (range: 6–20 months), contrasting the homogeneity of observed recommendations worldwide. Furthermore, we show that the optimal age depends on the local epidemiology of measles, with a lower optimal age predicted in populations suffering higher transmission. Overall, our results suggest the scope for public health authorities to tailor the recommended schedule for better measles control.

## Introduction

Measles is a highly contagious childhood infection^1^ caused by the measles virus. The virus is primarily spread through respiratory droplets and aerosols^2^, and symptoms include cough, fever, malaise, and a characteristic maculopapular rash^1^. Historically, measles was a major childhood disease, infecting almost all individuals in early life^3^ and resulting in 2–3 million deaths per year^1^. The introduction of measles vaccines in the 1960s significantly reduced the global number of measles cases and deaths^4^, with estimated deaths in 2021 reduced to approximately 128,000^5^.

However, despite the indisputable global success of the vaccine, measles remains endemic in multiple countries. Many regional elimination targets for 2020 were not met^6^, reflecting the difficulty of reaching the high immunization coverage needed for measles elimination^4^. These difficulties were compounded during the COVID-19 pandemic, which caused interruptions in routine vaccinations and supplementary immunization activities (SIAs)^6,7^, resulting in new measles outbreaks in 18 countries^6^. Since 2020, 140 countries have reported at least 1 case per year to the World Health Organization (WHO), and over 30 countries have reported over 1,000 cases in a year^8^.

Although the immunity gaps that drive continued measles transmission are mainly caused by insufficient vaccine coverage, they also result from vaccine failures. One avertable cause of these vaccine failures is the vaccination age: vaccination with the first dose of measles-containing vaccine (MCV1) at younger ages results in a higher risk of vaccine failure^9^. Two main mechanisms have been proposed to explain this result: blunting by maternal antibodies and immaturity of the infant immune system^10,11^. However, despite the potential consequences on measles control – e.g., changing the recommended MCV1 age as a potential control intervention – the impact of vaccination age on vaccine effectiveness (VE) has only recently been quantified^9^.

As illustrated in Figure 1a, the increasing effectiveness of MCV1 with age should result in a trade-off in risks when recommending MCV1 age: reducing the age of vaccination increases the risk of vaccine failure, while increasing the age worsens the risk of infection before vaccination. Hence, by balancing these risks, the recommended MCV1 age may be optimized to minimize measles incidence. Furthermore, location-specific factors, such as transmission level, are expected to affect this trade-off by changing the mean age of infection (MAI), resulting in different optimal ages^12,13^. Following this conceptual model, one expects the optimal vaccination age to depend on the local epidemiology of measles.

**Figure 1:**
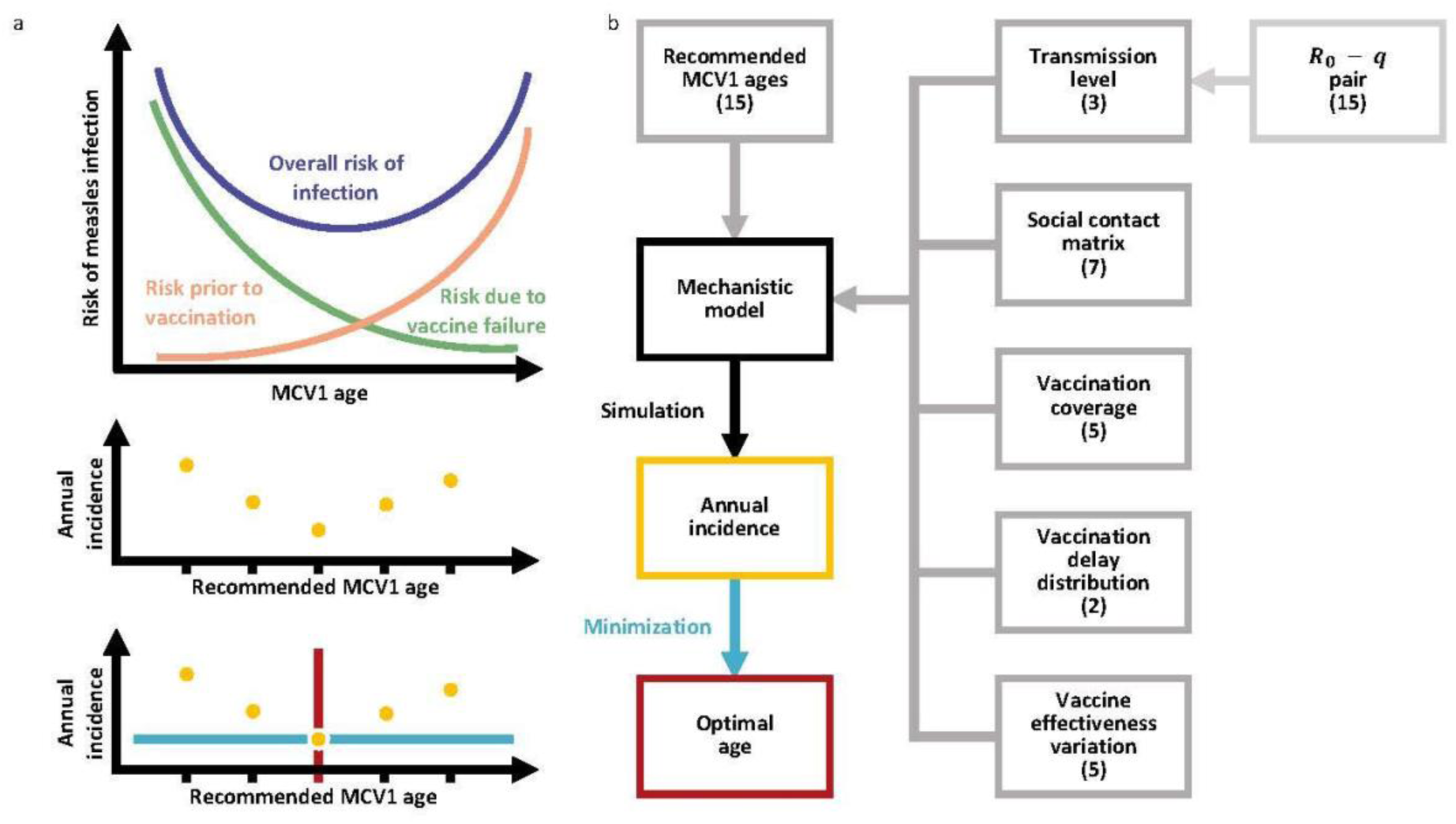
Trade-off in risks and resulting framework for calculating the optimal age to recommend MCV1. a) Illustration of the risk trade-off that should be balanced when recommending MCV1 age. b) Conceptual framework for calculating the optimal age to recommend MCV1. Gray boxes indicate variables used to parameterize the mechanistic model of measles transmission and vaccination. Values in brackets indicate the number of variants used. The yellow dots and box reflect incidence, the blue arrow and line the minimization of the incidence, and red indicates the age at minimum incidence, the optimal age.

As seen in Figure 2, however, the global homogeneity in recommended MCV1 ages contrasts with the observed heterogeneity in measles incidence^14^. The partial Spearman rank correlation coefficient between MCV1 ages and countries’ mean annual incidence was only –0.12 (p-value: 0.53) in countries with ≥1 measles case per 1 million per year, when controlling for Human Development Index (HDI), an aggregate measure of countries’ longevity, education, and standard of living^15^. Of the 205 MCV1 recommendations obtained, 70% of recommendations were at 9 months (69 countries) or 12 months (109 countries), reflecting the recommendations from the WHO: MCV1 at 9 months in countries with ongoing transmission and at 12 months in countries with low transmission^14^. Although these recommendations generally reflect the trade-off in risks, they may not capture the complexity of factors that impact measles epidemiology, such as further transmission variation driven by differences in social contact patterns or vaccination coverage.

**Figure 2:**
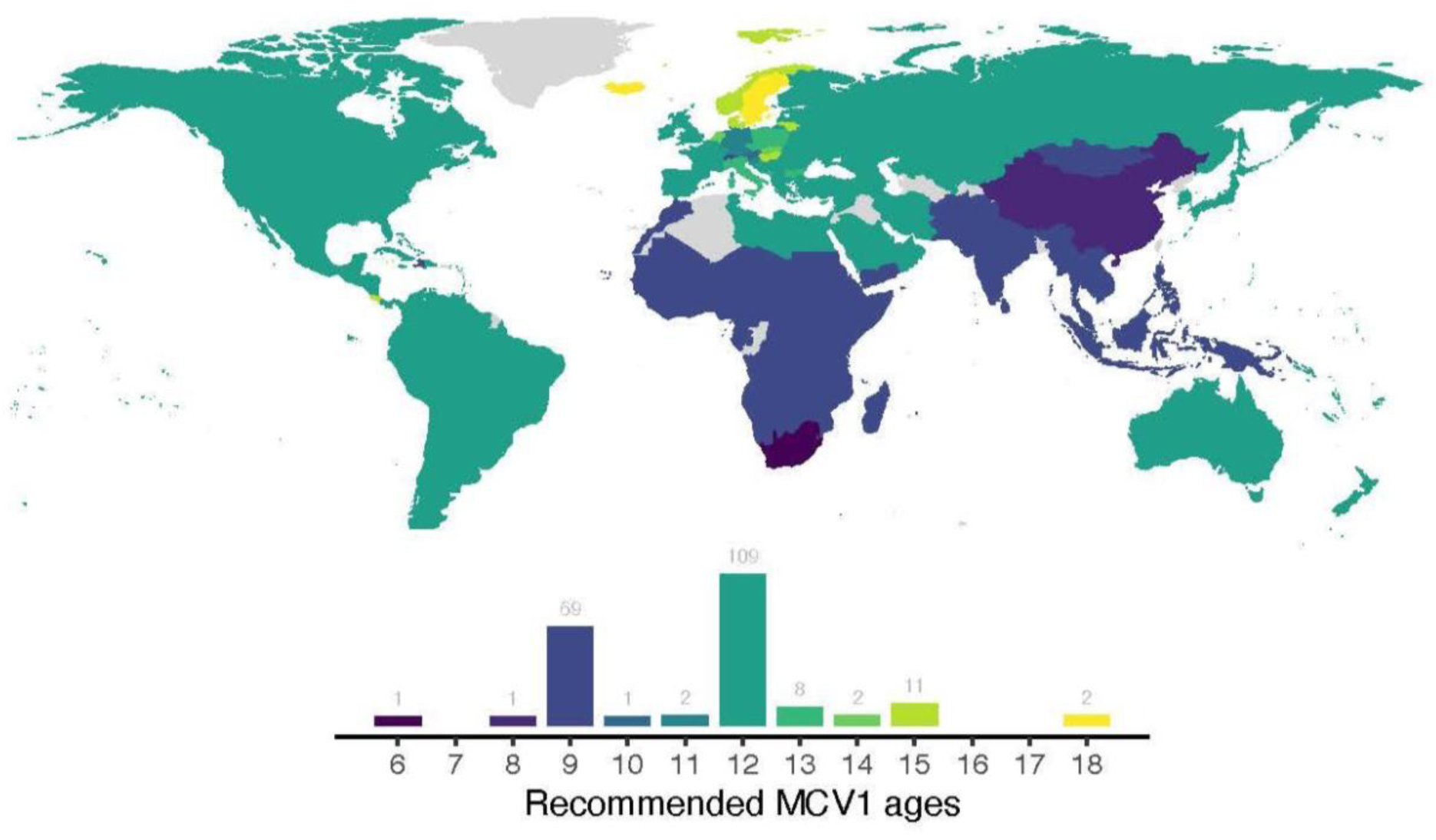
Map and distribution of recommended MCV1 ages. Reported recommended MCV1 ages by country, from the WHO^16^, ECDC^17^, and country-based reporting^18–23^. The histogram indicates the number of countries recommending MCV1 vaccination at each age.

The relative homogeneity in MCV1 recommendations suggests opportunities for refining the MCV1 age to leverage this risk trade-off. Here, we propose a new method—based on a mechanistic model of endemic measles transmission incorporating realistic, data-driven models of MCV1 delay and VE variation with age—to estimate the optimal age to recommend MCV1. As a proof of concept, we used this method to estimate the optimal MCV1 age in a range of synthetic test populations. In these populations, we varied parameter values across realistic ranges to identify factors determining optimal age (Figure 1b). We show that a mismatch between the optimal and recommended ages can potentially increase measles incidence. Furthermore, we show that the optimal age is sensitive to location-specific determinants of measles epidemiology, with transmission level having the greatest effect, followed by the social contact structure and vaccination coverage. Overall, our study suggests that the optimal MCV1 age is highly population-specific and, hence, more heterogeneous than the current recommendations reflect. Our findings thus suggest the potential to adjust MCV1 ages to reduce measles incidence, taking steps toward eventual elimination.

## Methods

### Data on measles incidence, recommended MCV1 age, and the Human Development Index

We gathered data on MCV1 recommended age from the WHO^16^ and the European Centre for Disease Prevention and Control (ECDC)^17^, supplemented with reports from individual countries in cases of missing data^18–23^. We also collected country-level estimates of annual measles incidence for 2010–2019 from the WHO^12^ and HDI values for the same period from the United Nations Development Program^15^.

### The relationship between MCV age and vaccine effectiveness

To quantify how vaccination age affects MCV VE, we fitted a statistical model to reported estimates of MCV VE, obtained from a systematic review^9^. For MCV1 VE, to reduce uncertainty in MCV1 age, we only included estimates with an MCV1 age interval of <3 months. For the included MCV1 VE estimates, we calculated each estimate’s standard error from reported 95% confidence intervals^24^.

We then fitted a shape-constrained additive model (SCAM)^25^ to the logit-transformed MCV1 VE estimates. Specifically, we used a monotonically increasing P-spline basis with 4 knots, weighted according to precision, with standard errors transformed using the Delta method. We then calculated approximate simultaneous confidence intervals to assess uncertainty in model fit^26^. To include this uncertainty in the transmission model, we considered 5 curves corresponding to the predicted 2.5%, 25%, 75%, and 97.5% quantiles and the maximum likelihood estimate (MLE) from the SCAM.

For MCV dose 2 (MCV2) VE, too few estimates were available to assess age dependencies (Supplementary Figure 1). We, therefore, assumed MCV2 VE to be constant and equal to the mean MCV2 VE.

### The distribution of MCV1 delay

To incorporate realistic distributions of MCV delay (*i.e.*, the delay between the recommended age and the actual age of administration), we obtained data on MCV1 delay in 45 low and middle-income countries (LMICs)^27^, where most measles deaths occur^5^. The data consisted of the observed 25%, 50%, and 75% quantiles of the delay distribution. We excluded any countries with negative median delay. We initially fitted an Exponential distribution, which failed to capture the observed long right tails. Hence, we then fitted a Lomax distribution^28^ (an extension of the Exponential distribution with longer right tails to capture long delays) to every country by minimizing the squared distance between the simulated and observed quantiles. To summarize the variation in delay distributions, we clustered the Lomax distribution parameters using Partitioning Around Medoids (PAM) Euclidean distance clustering^29^. The number of clusters was determined using the average silhouette method^30^ and the Gap statistic^31^, with 500 bootstraps.

### Model of measles transmission and vaccination

To simulate measles incidence when recommending MCV1 at different ages, we constructed a mechanistic model of measles transmission and vaccination, incorporating the aforementioned data-driven statistical models of MCV1 delay and age-specific MCV1 VE. The model was a deterministic SIR model, which split the population by infection status into susceptible, infectious, recovered, and protected by maternal antibodies. For sufficiently large populations, deterministic models have been shown to capture the dynamics of measles^32,33^.

The model was age-structured, to allow the vaccination age to vary. The model split the population into monthly age groups between ages 0 to 59 months, then into 5-year age groups between ages 5 to 79. We assumed a uniform age distribution and constant population size. Contacts between age groups were parameterized using data-derived social contact matrices (SCM)^34,35^. To capture the variability in social contact structure, we selected 7 SCMs derived from China, India, Japan, Moscow, South Africa, the UK, and the USA, representing the clusters identified in a previous study that clustered SCMs from 35 countries and 277 subnational administrative regions^34^.

To model vaccination with two doses of MCV, we added a vaccinated susceptible state to model infants with primary vaccine failure (*i.e.*, infants who received the vaccine but failed to mount an effective immune response^36^). Vaccination was assumed to occur when aging from one age group to the next. For the first dose (MCV1), at a given age, individuals were either vaccinated or not vaccinated, determined by the recommended MCV1 age, delay distribution, and MCV1 coverage. If unvaccinated, individuals entered the next age group’s susceptible compartment. If vaccinated, the probability of successful vaccination was determined by the VE-age relationship. If successful, infants were protected and entered the recovered compartment of the next age group. If unsuccessful, they remained unprotected and entered the next age group’s vaccinated-susceptible compartment. The process remained the same for MCV2, but vaccination occurred when aging from the vaccinated susceptible compartment. The recommended MCV2 age was modeled as 6 months after the recommended MCV1 age, aligning with the modal gap between reported MCV schedules^16–23^.

Full model details, including parameterization, are included in the supplementary materials (Supplementary Table 1).

### Recapitulating reported pre-vaccine mean ages of measles infection

To calibrate transmission parameters, we compared the simulated MAI with historical reports from the pre-vaccine era^37^, which ranged from 24 months to 72 months (Supplementary Table 2). We grouped the MAI into three transmission levels: 48–72 months (low-transmission level), 36–48 months (medium-transmission level), and 24–36 months (high-transmission level). Based on the evidence of age heterogeneities in transmissibility^38,39^, we incorporated a parameter (*q*) representing the transmissibility of <5-year-olds relative to ≥5-year-olds. Finally, for a range of fixed *R*_0_ values in the interval 10–20, we calibrated *q* by fitting the predicted pre-vaccine MAI to 3 MAI target values (lower bound, mid-value, and upper bound) in each transmission level. For each target MAI, 5 *R*_0_—*q* pairs were selected, resulting in a total of 15 pairs for each transmission level.

The model was run assuming a constant population of 10 million for 500 years, at which point convergence to the equilibrium solution was determined by the magnitude of the derivatives^40^. If this convergence criterion was not fulfilled, the final 20 years of the simulation were extracted, and a linear model fit to the modeled cases. The simulation was judged to have converged if the slope of the linear model was less than 10^−3^ per day, corresponding to a change of <1 case per year. If convergence was achieved, the modeled MAI was calculated and compared against the target MAI using the sum of squares.

### The optimal age to recommend MCV1

We simulated recommending MCV1 at different ages, monthly from 6 to 20 months. For each recommended age, we calculated the corresponding annual incidence at equilibrium, then identified the MCV1 age that minimized the incidence aggregated over all age groups. Furthermore, to identify factors that have the greatest impact on the optimal age we varied these factors across realistic values (Figure 1b).

We simulated measles annual incidence using the model described above, simulating without vaccination for 50 years, then introducing vaccination and running the model for a further 950 years. Vaccination was modeled as beginning from the recommended MCV1 age, with delays in MCV1 and MCV2 following the delay distributions described above. The vaccine coverage was defined as the proportion of a birth cohort vaccinated by 24 months after the recommended MCV dose age. Based on reported MCV coverages from the WHO^14^, we set MCV1 coverage at 45%, 55%, 65%, 75%, and 85%, and MCV2 coverage at 5% points lower than the set MCV1 coverage.

To assess variation in optimal ages, we estimated the optimal age for every combination of SCM (China, India, Japan, Moscow, South Africa, UK, USA), vaccine coverage (MCV1 coverages: 45%, 55%, 65%, 75%, 85%), delay distribution (short delay and long delay), VE curve (2.5%, 25%, 75%, 97.5% quantiles, and the MLE), and transmission level (low-, medium-, and high-transmission, with 15 *R*_0_–*q* pairs for each level), see Figure 1b. Any combination that failed to converge to the equilibrium solution according to the abovementioned convergence criteria, at any recommended MCV1 age, was removed. Optimal ages were then calculated and compared. To facilitate this comparison and evaluate the current WHO clustering of recommendations, we clustered (using PAM clustering^29^ based on Euclidean distance, and the silhouette method^30^ to determine cluster sizes) the estimated optimal ages to identify groups of SCM.

### Numerical Implementation

Analysis was carried out using R version 4.1.1^41^, using the R package “tidyverse”^42^. Partial correlations were calculated using the package “ppcor”^43^. SCAMs were fitted using the package “scam”^44^. Lomax distributions were fitted using the R package “optim”^45^, using the algorithm “L-BFGS-B”^46^. PAM clustering was carried out using the R packages “cluster”^47^ and “factoextra”^48^. The measles model was implemented in C and R, using the R package “pomp”^49^. Parameter fitting was carried out using the subplex algorithm^50^, in the R package “nloptr”^51^. Mixed-effect models were fitted using the package “lme4”^52^. Figures were created using the R packages “ggplot2”^53^, “viridis”^54^, “wesanderson”^55^, “patchwork”^56^, and “ggh4x”^57^.

## Results

### MCV1 effectiveness increases with age of receipt

After applying our inclusion criteria, we analyzed a total of 52 VE estimates from 16 studies. As shown in Figure 3a, the point estimates and the confidence intervals of VE varied greatly. Lower ages displayed particularly high variation in VE estimates. A large part of the overall variation was captured by the SCAM (59.3% of deviance explained), which estimated an increase in VE with age, confirming the results of the earlier meta-analysis^9^. The SCAM estimated the VE at 64.5% at 6 months, approaching 100% by 20 months. Hence, the model confirms that, for infants ≤20 months, the effectiveness of MCV1 increases with age of receipt.

**Figure 3:**
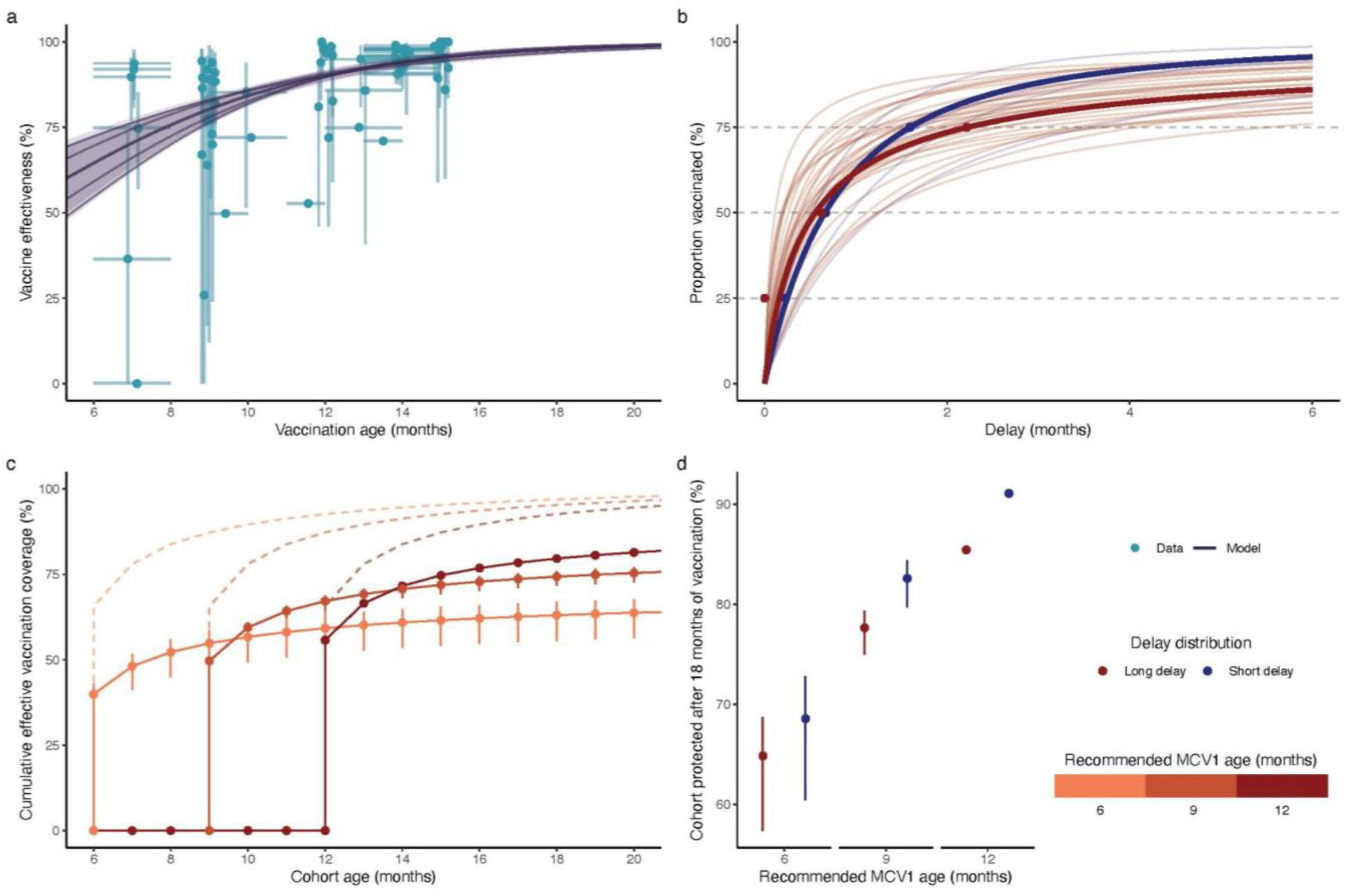
The impact of recommended MCV1 age on the effective vaccine coverage. a) MCV1 effectiveness with age of receipt. VE estimates are indicated with points, with vertical lines indicating the 95% confidence interval and horizontal lines indicating uncertainty in MCV1 age. The SCAM model is indicated in gray; the shaded area indicates the approximate simultaneous 95% confidence interval, and the lines indicate the 2.5%, 25%, MLE, 75%, and 97.5% quantiles. b) Cumulative delay distributions. Cluster medoids are bolded, with points showing the associated delay data. c) Cumulate effective MCV1 coverage when recommending MCV1 at 6, 9, and 12 months, with long delay distribution. Cumulative vaccine coverage after 24 months is set to 100%. Dashed lines indicate the MCV1 coverage, points indicate the effective MCV1 coverage, with vertical lines indicating the 95% confidence intervals. d) Cumulative effective MCV1 coverage at 18 months when recommending MCV1 at 6, 9, and 12 months, for long and short delay distributions. Points indicate MLE estimates, and vertical lines indicate the 95% confidence intervals.

### Empirical data on the reported age of vaccination reveal MCV1 is frequently delayed

Based on data from 43 countries that met our inclusion criteria, we found that delays in receiving MCV1 were prevalent (median (range) of median delay: 0.6 (0.1, 1.3) months), exceeding 3 months for 25% of infants in 9 countries (median (range) of 75% delay quantile: 1.8 (0.4, 5.6) months).

In 41 of the 43 countries, we successfully fitted the Lomax distribution (see Supplementary Figure 2), which recapitulated the 50% and 75% quantiles of the observed delay distributions. Using PAM clustering, we identified two broad groups of countries (Figure 2b, Supplementary Figure 2): one group with longer right tails (long delay, 35 countries, medoid country: Uganda, survey median (IQR): 0.6 (0, 2.2) months, model median (IQR): 0.6 (0.2, 2.2) months) and another group with shorter right tails (short delay, 5 countries, medoid country: Turkey, survey median (IQR): 0.7 (0.2, 1.6) months, model median (IQR): 0.7 (0.2, 1.6) months). For both clusters and all observed countries, the estimated parameters resulted in median delays in the range 0.1 to 1.3 months, corresponding to a median delay of up to 14% when MCV1 was recommended at 9 months. Taken together, this analysis indicates MCV1 is frequently delayed, with important implications for measles control by vaccination, modeling the transmission dynamics of measles, and estimating the optimal age of MCV1.

By combining the delay distribution with age-specific MCV1 effectiveness, we calculated the cumulative effective vaccine coverage, defined as the proportion of a birth cohort protected by the vaccine by a given age (Figure 2c). This effective coverage reflected the conceptual trade-off in risks outlined in Figure 1a: increasing the recommended MCV1 age left a birth cohort susceptible to infection for longer but also increased MCV1 effectiveness and, hence, the long-term proportion of the cohort protected. Furthermore, the delay distribution also determined the effective coverage, with longer delays resulting in lower proportions of a birth cohort protected 18 months after the recommended age (Figure 2d).

### Heterogeneity in transmissibility is necessary to recapitulate pre-vaccine reports of measles MAI

When comparing model-derived MAIs to historical estimates in the pre-vaccine era, we found that, for typically reported values of *R*_0_^58^ between 12 and 18, the model failed to recapitulate the MAIs for multiple SCMs (Supplementary Figure 5). However, once age-specific transmissibility was included, the transmission model could recapitulate all historical estimates of MAIs for all SCMs, except the Chinese SCM for a MAI of 24 months. All fitted values of *q* and *R*_0_converged according to the convergence criteria. These fitted pairs displayed a negative association, such that increases in *R*_0_ were compensated by decreases in *q*. Hence, this calibration allowed us to define parameter regions that reproduce each transmission level (Figure 4) for inclusion in the transmission model.

**Figure 4:**
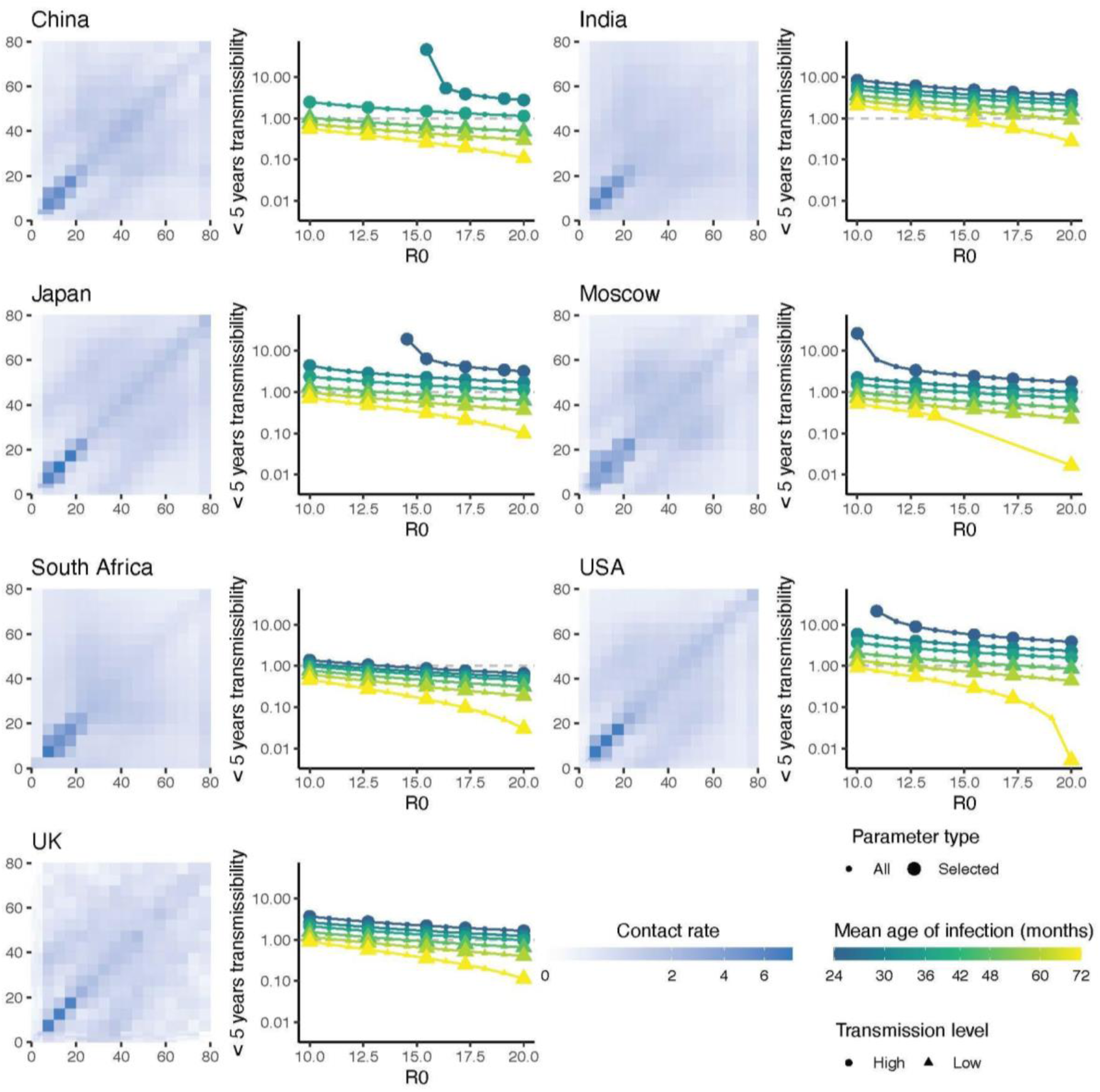
Basic reproductive number and relative <5 years transmissibility for 7 social contact matrices. For each SCM, the left-hand plots show the social contact rate between age groups. In the corresponding right-hand plots, points indicate the fitted values of *R*_0_ and *q* for each target MAI, with larger points indicating the values selected to parameterize the mechanistic model. Transmission levels are indicated by point shape. For clarity, medium-transmission level points are not shown.

### The optimal age to recommend MVC1 is sensitive to transmission level, contact structure, and vaccine coverage

Of all the parameter sets modeled, 99.4% fulfilled our convergence criteria. Across those, we identified a unique optimal age ranging between scenarios from 6 to 20 months. Furthermore, the predicted optimal ages varied greatly between scenarios (see Figure 5a-b). For example, the optimal age for the low-transmission level with the China SCM at 85% MCV1 coverage ranged from 15 to 20 months, whereas the high-transmission level at 45% vaccine coverage with the South Africa SCM ranged from 6 to 7 months. Moreover, recommending a non-optimal MCV1 age on incidence could result in up to a 2.6-fold increase in incidence, although the impact of an age mismatch was similarly scenario-dependent (see Figure 5a, Table 1).

**Figure 5:**
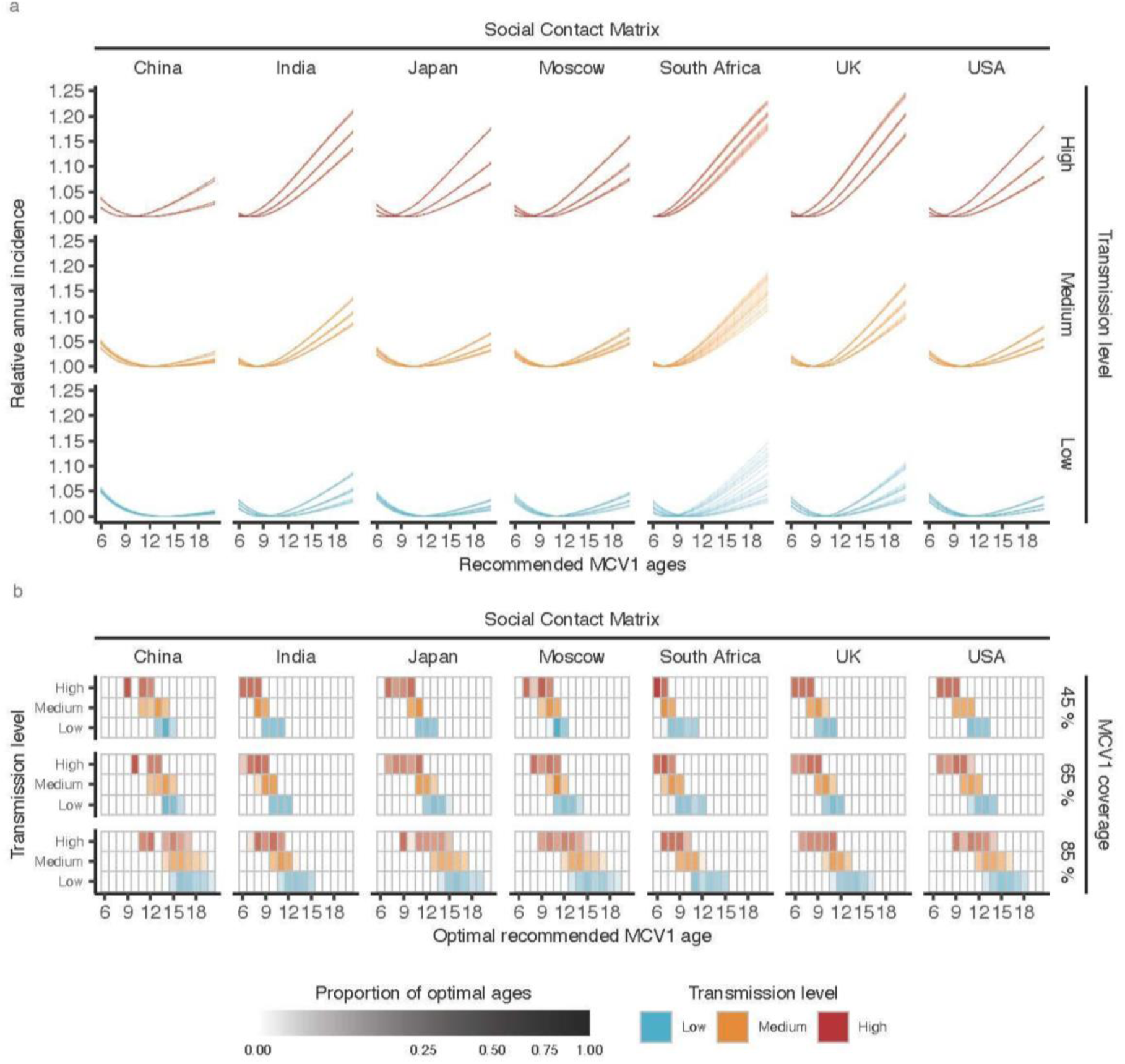
Estimating the optimal age to recommend measles vaccination. a) Estimated annual incidence when recommending MCV1 at ages 6–20 months, with 45% MCV1 coverage. Each line indicates the relative incidence for a given parameter set, relative to the minimum incidence for that parameter set. b) Heatmap of optimal ages. Opacity indicates the proportion of parameter sets with an optimum in a given MCV1 age. For clarity, the results for 55% and 75% vaccination coverage are not displayed.

**Table 1:**
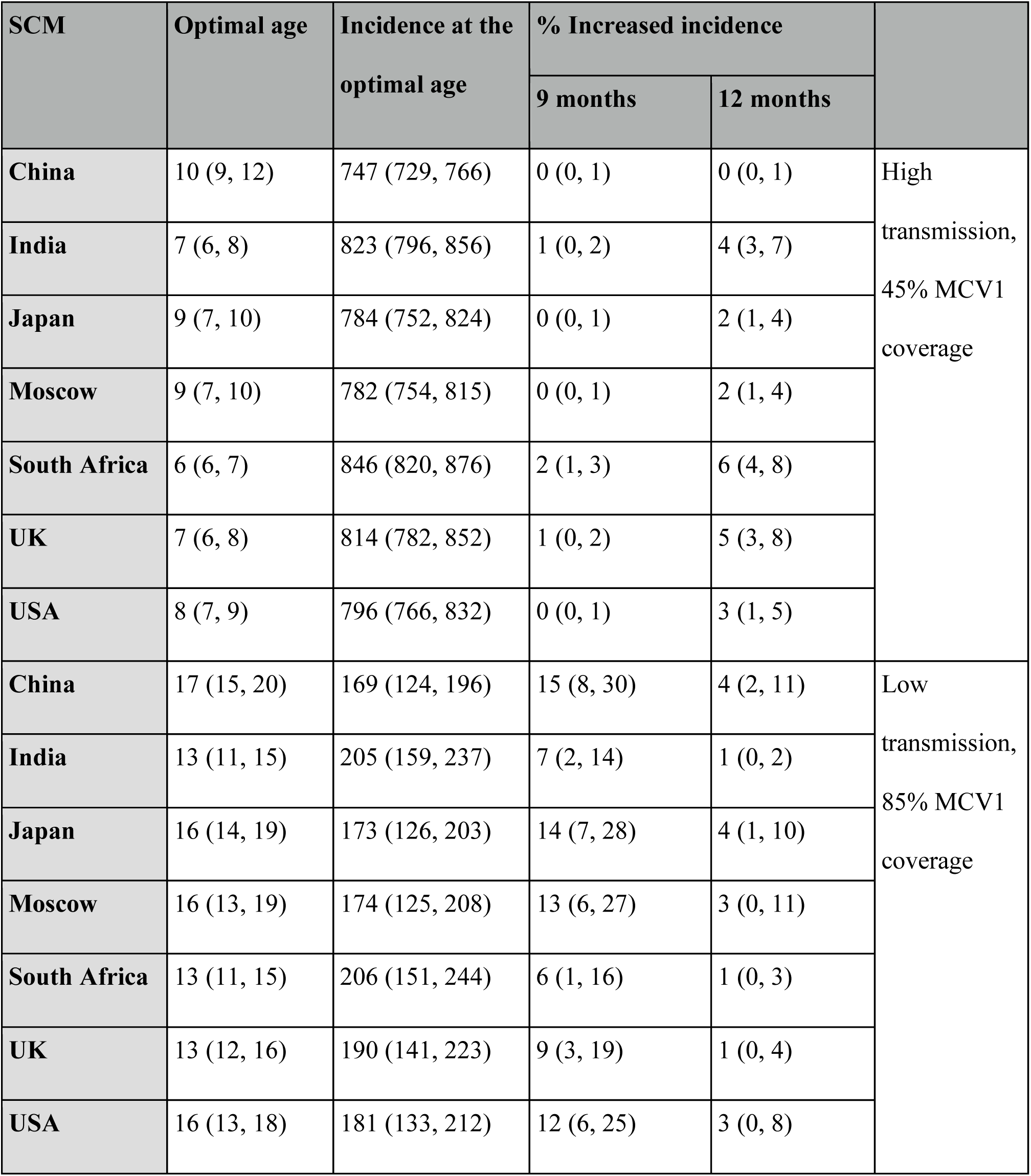
Estimated optimal ages and annual incidences. Optimal age indicates the mean (95% confidence interval) of the optimal ages from all parameter sets for the scenario. Incidence indicates the mean (95% confidence interval) annual incidence per 100,000 of the parameter sets at the optimal ages. Percentage increased incidence reflects the mean of the incidence increase compared to the optimal age incidence.

Of the factors we varied, the transmission level impacted the optimal age the most. At 45% MCV1 coverage, parameterized with the USA SCM, the optimal age ranged between 11 to 13 months in a low-transmission setting and 7 to 9 months in a high-transmission setting. More generally, increasing transmission from low to medium decreased the optimal age by an average of 1.6 months, and increasing from low to high transmission resulted in an average decrease of 3.6 months (Supplementary Table 3). Higher transmission resulted in lower MAIs and increased the risk of infection at younger ages, thus resulting in younger optimal ages to compensate for this risk. Accordingly, increases in the pre-vaccine MAIs resulted in increases in the optimal age (Supplementary Figure 6a). After controlling for the MAI, the additional impact of *R*_0_ and *q* on the optimal age was minimal (data not shown).

Social contact structure also affected the optimal age to recommend MCV1. Even after controlling for transmission level and vaccine coverage, the range of optimal ages varied between SCMs: for example, from 6 to 7 months for the South Africa SCM to 9 to 12 months for the China SCM for a scenario with high transmission at 45% coverage. Depending on the MCV coverage–transmission level scenario considered, the optimal ages clustered into 2–5 groups. However, ≥3 groups were typically required to capture the heterogeneity in optimal ages between SCMs (13/15 scenarios, Supplementary Figure 7). In most scenarios, the China SCM and the South Africa SCM tended to cluster independently, representing the SCMs with the oldest and youngest optimal ages respectively, with other SCMs clustering together, with optimal ages between these groups.

Finally, vaccine coverage also impacted the optimal age. Specifically, increased coverage reduced measles incidence, resulting in higher optimal ages. For example, optimal MCV1 ages in a low-transmission setting with the South Africa SCM ranged from 7 to 11 months at 45% vaccine coverage and from 11 to 15 months at 85% vaccine coverage. In general, when accounting for transmission level and SCM, a 10-percentage point increase in MCV1 coverage resulted in an average increase in optimal age of 0.8 months (Supplementary Table 4). Overall, these results demonstrate the importance of location-specific factors of measles epidemiology for vaccine policy.

### The impact of variations in age-specific MCV1 effectiveness and delay distribution on the optimal age is minor

Uncertainty in the vaccine effectiveness curve only marginally affected the optimal age. When holding all other parameters constant, varying the curve resulted in changes in the optimal age in only 12.0% of parameter sets. In cases where the optimal age varied, the effect was inconsistent, but generally, as the quantile of the VE curve increased, the optimal age decreased (Supplementary Figure 6b).

Uncertainty in the MCV delay distribution had a similarly minor impact on optimal age. When holding all other parameters fixed, changing the delay distribution resulted in a change in optimal age in only 14.6% of parameter sets. When changes occurred, a higher optimal age was predicted for the short delay distribution (Supplementary Figure 6c).

## Discussion

In this study, we developed a new method, based on a mechanistic model of measles transmission and vaccination, to estimate the optimal age to recommend MCV1. In particular, this model captured several complexities of measles epidemiology, including age-specific contacts, vaccination delays, and VE variations with age of receipt. For every scenario tested, we could identify a unique optimal age in the range of 6–20 months, contrasting the two ages recommended by the WHO. Moreover, we found that the optimal age was governed by location-specific factors, namely transmission level, vaccine coverage, and social contact structure. Overall, our results suggest that, in addition to increasing vaccine coverage, adjusting the recommended vaccination age could help minimize immunity gaps and reduce measles incidence, taking steps toward eventual elimination from endemic settings.

A key result from our study is that the optimal age to recommend MCV1 may depend on the local epidemiology of measles. Specifically, we predict that populations suffering from higher measles transmission require a lower vaccination age. More generally, the impact of certain factors of local epidemiology can be understood by considering their effect on the MAI and transmission after vaccine introduction. This explains the effect of pre-vaccination transmission levels (controlled by the parameters *R*_0_ and *q*), which are correlated with post-vaccination transmission levels. Similarly, increasing vaccination coverage is dynamically equivalent to reducing transmission^59^, hence resulting in higher optimal ages. However, even at fixed transmission levels, the impact of social contact structure was strong. This result shows that, in addition to broad metrics quantifying the transmission of measles, detailed knowledge of social contact structure, quantified by data-derived SCMs, is needed to identify the optimal in a given population.

In our simulations, two transmission parameters were required to recapitulate the range of transmission intensities (quantified by the MAI) observed in the pre-vaccine era. Unlike earlier modeling studies^60,61^, varying only the basic reproduction number was insufficient to reach target MAIs for all contact matrices. This discrepancy may be explained by the high age resolution and the inclusion of realistic SCMs in our model. More generally, this result suggests age heterogeneities beyond social contacts are necessary for the design of realistic models of measles. Here, we allowed the relative transmissibility of <5 year olds to vary, and calibrated values varied across multiple orders of magnitude (range: 0.005–46.3). This heterogeneity could also be interpreted as a correction to the SCM, as the SCMs used were derived for more modern populations than the pre-vaccine MAI estimates.

As the main goal of our study was to establish a proof of concept, we chose the minimization endpoint of equilibrium incidence to estimate the optimal age in endemic settings. In real-world applications, however, this endpoint should be defined over shorter time scales, reflecting the time frame of control, and should be reassessed frequently as the optimal ages change with decreasing transmission. Additionally, other endpoints like hospitalization or deaths may be considered but will require extending our model to represent additional mechanisms of vaccine protection, such as reduced disease severity in vaccine-breakthrough cases^62^. Importantly, considering other endpoints may change the nature of the trade-off in risks, in particular because of the increased incentive to vaccinate early if mortality is increased in younger ages^63^. Similarly, other endpoints, like the risk of invasion, will be needed to estimate the optimal age in elimination settings, where vulnerability to outbreaks may persist due to residual pockets of susceptible individuals^64^. Applying our model in such settings will require a stochastic formulation, due to the low number of cases and frequent extinctions that deterministic models cannot capture well.

Furthermore, the real-world application of our method will require additional components beyond population-specific information on the SCM, vaccine coverage, and delay distribution, to fully characterize measles epidemiological dynamics in a target population. These include demography, as changes in population structure are expected to affect age-specific transmission dynamics^65^, with more circulation expected in younger populations^66^. A second key component is seasonality in transmission, which can result from term-time increases in contacts among school-aged children^67^ or the effect of climate on virus transmissibility^68^. Therefore, a prerequisite to applying our method is a detailed model—identified, for example, by fitting to long-term incidence data using modern statistical inference techniques^49^—for capturing the local drivers of measles transmission.

Another key consideration when applying our proposed method is SIAs. These additional immunization campaigns aim to rapidly increase population immunity by vaccinating target demographics—typically children aged ≤14 years—regardless of vaccination history^69^. In general, such campaigns are expected to reduce transmission and, thus, increase the optimal age for MCV1. Hence, in settings where SIAs are routinely administered, MCV1 age should be optimized to maximize the effect of both interventions.

Beyond the components listed above, our method could be extended to consider the effects of MCVs on other pathogens. Indeed, measles infection can cause immune amnesia, whereby the suppression of immune cells partially erases immune memory to previously encountered pathogens^70^. As a result, MCVs have beneficial indirect effects on other infectious diseases^71^. In addition, it has been proposed that MCVs directly affect non-measles pathogens, perhaps because of enhanced trained immunity^72^. Beyond MCVs, our proposed method could also be applied to estimate the optimal age for vaccines against other childhood infections. We expect the relationship between vaccination age and VE to be qualitatively similar for other vaccines, even though the empirical evidence has remained more limited than for measles^73,74^. As the measles vaccine is often combined with the mumps and rubella vaccines (MMR), the natural next candidates would be these two vaccines. Because mumps and rubella infection have different transmissibility than measles (Mumps *R*_0_: 4–7^75^, Rubella *R*_0_: 6–7^75^), the risk trade-off underlying the optimal age is expected to differ. Hence, a future research question is how to extend our approach to identify the optimal age for combined vaccines.

Despite the availability of effective vaccines for over 60 years, measles remains a considerable threat in many countries. Here, we propose that, alongside ever-necessary efforts to increase vaccine coverage, another effective intervention to reduce measles cases may be to tailor the vaccination age. Hence, our results suggest the scope for public health authorities to improve measles control and reach for eventual elimination by customizing the recommended vaccination schedule. More generally, as the trade-off underlying the optimal age is not specific to measles, our results could have ramifications for controlling many other vaccine-preventable diseases.

## Data availability / Code availability

Code will be deposited in the Open Research Data Repository of the Max Planck Society, Edmond (https://edmond.mpg.de), before publishing.

## Acknowledgements

This work was supported by the Max Planck Society (Berlin, Germany). Computations were performed at the Max Planck Computing and Data Facility (MPCDF). We thank Professor Pejman Rohani for his valuable feedback on the manuscript.

## Author contributions

EG and MDdC conceptualized the project, and designed the methods. Implementation was carried out by EG, and reviewed by LABG. EG and MDdC wrote the manuscript, with support from MB and LABG. MDdC supervised the project.

