## Supplementary materials for "Estimating the optimal age for infant measles vaccination"

**Supplementary material**

Elizabeth Goult[1,*] Laura Andrea Barrerro Guevara[1,2]
Michael Briga[1] Matthieu Domenech de Cellès[1]

1. Infectious Disease Epidemiology group, Max Planck Institute for Infection Biology, Charitéplatz 1, Campus Charité Mitte, 10117, Berlin, Germany
2. Institute of Public Health, Charité—Universitätsmedizin Berlin, 10117 Berlin, Germany

**2 Supplementary tables** **7**

**3 Supplementary figures** **9**

### 1 Description of the model of measles transmission and vaccination

The measles model split the modeled population by disease state into susceptible ($S$), infected ($I$), recovered ($R$), and protected by maternal antibodies ($M$), along with a vaccinated-susceptible ($VS$) compartment to model people who received at least one dose of measles containing vaccine (MCV), but failed to mount a protective immune response. The total population was hence defined as $N = S + I + R + M + VS$.

The model was also age structured, splitting each disease state into age groups, resulting in $M_i$, $S_i$, $I_i$, $R_i$, and $VS_i$, for each age group $i$. Hence, the total population in age group $i$ was defined as $N_i = M_i + S_i + I_i + R_i + VS_i$. People aged out of each age group at a rate $\delta_i$, determined by the width of the age group. To facilitate the analysis, the population was split into monthly age groups between 0–59 months, then 5 yearly from 5 to 79 years, resulting in 75 age groups. As shown in Supplementary Figure 3, we modeled vaccination as occurring when populations transition between age groups

#### 1.1 Protected by maternal antibodies

Infants were born into $M_1$ at a rate $pb$, where $b$ is the birth rate, and $p$ is the proportion of infants born to protected mothers. Hence, $p$ represents the prevalence of women of childbearing age who were recovered or successfully vaccinated against measles.

$$p = \frac{\sum_{i=63}^{i=71} R_i}{\sum_{i=63}^{i=71} N_i}. \tag{1}$$

Infants leave the compartment either when maternal antibody protection wanes at rate $m$, or when they age out. Hence, the equation describing the dynamics of the population protected by maternal antibodies compartment is

$$\frac{\mathrm{d}M_1}{\mathrm{d}t} = pb - mM_1 - \delta_1 M_1, \tag{2}$$

for the first age group, and,

$$\frac{\mathrm{d}M_i}{\mathrm{d}t} = \delta_{i-1} M_{i-1} - mM_i - \delta_i M_i, \tag{3}$$

for all older age groups.

#### 1.2 Susceptible

Infants born to unprotected mothers entered the first susceptible compartment at rate $(1 - p)b$. People also entered susceptible compartments by losing maternal antibodies, and, for $i > 1$ by aging into the susceptible compartment from the age group before without being vaccinated. People then left the susceptible compartment either by aging out, or by being infected.

We modeled vaccination occurring at the transitions between age groups, hence, vaccination with MCV dose 1 (MCV1) occurred with probability $v_i^{(1)}$, and was successful with probability $VE_i^{(1)}$ (the age dependent MCV1 vaccine effectiveness (VE)) and unsuccessful with probability $1 - VE_i^{(1)}$. The remaining infants aged into the next age group's susceptible compartment.

The probability of MCV1 vaccination is determined by the MCV1 delay distribution and overall MCV1 coverage, and is equivalent to the probability of receiving MCV1 at age $i$ given MCV1 was not received at any prior age.

##### 1.2.1 Incorporating vaccination

Let $V_i^{(k)}$ be the event that vaccination with MCV dose $k$ occurs at age $i$, and $V^{(k)}$ the event that vaccination with MCV dose $k$ occurs at all. Then, the delay distribution and the initial vaccination age ($i_0^{(k)}$) for MCV dose $k$ gives the probability of vaccination with MCV dose $k$ occurring at a given age $i$, given vaccination with MCV dose $k$ occurs at all, $p_i^{(k)}$. Hence,

$$p_i^{(k)} = Pr(V_i^{(k)}|V^{(k)}). \tag{4}$$

We assume that vaccination with dose $k$ occurs within 24 months of $i_0^{(k)}$. Hence, the probability that vaccination with MCV dose $k$ occurs at age $i$ becomes

$$\bar{p}_i^{(k)} = Pr(V_i^k) = \frac{\kappa_k p_i^{(k)}}{\sum_{i=i_0^{(k)}}^{i_0^{(k)}+24} p_i^{(k)}}, \tag{5}$$

where $\kappa_k$ is the dose dependent MCV coverage, $Pr(V^{(k)})$.

For parameterization of our transmission model, we require $v_i^{(k)}$, the probability that vaccination with MCV dose $k$ occurs at age $i$ given that vaccination does not occur at ages $i_0^{(k)}$ to $i-1$. Let $!(V_{i0:i-1}^{(k)})$ be the event that vaccination with dose $k$ does not occur between ages $i_0^{(k)}$ and $i-1$. Hence, we require

$$v_i^k = Pr(V_i^k|!(V_{i0:i-1}^{(k)})). \tag{6}$$

By Bayes' theorem, this becomes

$$v_i^{(k)} = \frac{Pr(!(V_{i0:i-1}^{(k)})|V_i^k)Pr(V_i^k)}{Pr(!(V_{i0:i-1}^{(k)}))}. \tag{7}$$

As $Pr(!(V_{i0:i-1}^{(k)}))$ is the probability of not being vaccinated by age $i-1$, this is equivalent to $1 - \sum_{i=i_0}^{i-1} \bar{p}_i^{(k)}$. Hence,

$$v_i^{(k)} = \frac{\bar{p}_i^{(k)}}{1 - \sum_{i=i_0}^{i-1} \bar{p}_i^{(k)}}. \tag{8}$$

##### 1.2.2 Susceptible equations

Taken together, the equations for the susceptible compartments become

$$\frac{\mathrm{d}S_1}{\mathrm{d}t} = (1-p)b + mM_1 - \lambda_1 S_1 - \delta_1 S_1, \tag{9}$$

for the first age group, and

$$\frac{\mathrm{d}S_i}{\mathrm{d}t} = mM_i + \delta_{i-1}(1 - v_{i-1}^{(1)})S_i - \lambda_i S_i - \delta_i S_i, \tag{10}$$

for older age groups.

Here, the term $\lambda_i S_i$ models infection, where the age-dependent force of infection is

$$\lambda_i = \beta\big(q \sum_{j=1}^{60} \frac{c_{i,j} I_j}{N_j} + \sum_{j=61}^{75} \frac{c_{i,j} I_j}{N_j}\big). \tag{11}$$

$\beta$ denotes the infection probability given contact, derived from the basic reproduction number $R_0$ using the next-generation matrix [1], $q$ denotes the relative transmissibility of the under 5 years age group, and $c_{i,j}$ the per-capita frequency of contact between age groups $i$ and $j$.

#### 1.3 Infected

People entered the infected compartment for a given age group either through infection from the susceptible compartment or the vaccinated-susceptible compartment in the same age group, or by aging into the compartment from the previous age group's infected compartment. People exited the compartment by recovering from measles at rate $\gamma$, or by aging out of the compartment. Hence, the equations describing the dynamics of the infected populations are

$$\frac{\mathrm{d}I_1}{\mathrm{d}t} = \lambda_1(S_1 + VS_1) - \gamma I_i - \delta_1 I_1, \tag{12}$$

for the first age group, and

$$\frac{\mathrm{d}I_i}{\mathrm{d}t} = \lambda_i(S_i + VS_1) + \delta_{i-1} I_{i-1} - \gamma I_i - \delta_i I_i, \tag{13}$$

for all other age groups.

#### 1.4 Recovered

Individuals entered the recovered compartment or a given age group via 4 paths: by recovering from measles infection; by aging out of the previous age groups recovered compartment; by being successfully vaccinated with MCV1, occurring with rate $\delta_{i-1} v_{i-1}^{(1)} VE_{i-1}^{(1)}$; or, by being successfully vaccinated with MCV dose 2 (MCV2), following unsuccessful vaccination with MCV1, with rate $\delta_{i-1} v_{i-1}^{(2)} VE_{i-1}^{(2)}$. Recovered individuals then remained in the recovered compartment and age into the next age group. The equations modeling the dynamics are hence,

$$\frac{\mathrm{d}R_1}{\mathrm{d}t} = \gamma I_1 - \delta_1 R_1, \tag{14}$$

for age group 1, and

$$\frac{\mathrm{d}R_i}{\mathrm{d}t} = \gamma I_i + \delta_{i-1}R_{i-1} + \delta_{i-1}v_{i-1}^{(1)}VE_{i-1}^{(1)}S_{i-1} + \delta_{i-1}v_{i-1}^{(2)}VE_{i-1}^{(2)}VS_{i-1} - \delta_i R_i, \tag{15}$$

for all older age groups.

#### 1.5 Vaccinated susceptible

Individuals entered an age group's vaccinated susceptible compartment either through unsuccessful vaccination with MCV1 with rate $\delta_{i-1}v_{i-1}^{(1)}(1-VE_{i-1}^{(1)})$, through unsuccessful vaccination with MCV2 with rate $\delta_{i-1}v_{i-1}^{(2)}(1-VE_{i-1}^{(2)})$, or by aging out of the prior vaccinated susceptible compartment without receiving MVC2, with rate $\delta_{i-1}(1-v_{i-1}^{(2)})$. People left the compartment either by being infected, by aging out without receiving MVC2, by receiving unsuccessful MCV2 vaccination, or receiving successful MCV2 with rate $\delta_i v_i^{(2)}VE_i^{(2)}$. Hence, the equation for the first vaccinated susceptible age group is

$$\frac{\mathrm{d}VS_1}{\mathrm{d}t} = -\lambda_1 VS_1 - \delta_1 VS_1, \tag{16}$$

and,

$$\frac{\mathrm{d}VS_i}{\mathrm{d}t} = \delta_{i-1}v_{i-1}^{(1)}(1-VE_{i-1}^{(1)})S_{i-1} + \delta_{i-1}(1-v_{i-1}^{(2)}VE_{i-1}^{(2)})VS_{i-1} - \lambda_i VS_i - \delta_i VS_i, \tag{17}$$

for all older age groups.

#### 2 Supplementary tables

| Parameter | Interpretation | Value | Source |
|---|---|---|---|
| $i$ | Age group | 1–75 | - |
| $N$ | Total population size | 10 million | - |
| $\delta_i$ | Age-specific aging rate | 0.0005, 0.03 day$^{-1}$ | - |
| $p$ | Proportion of infants born to protected mothers | Equation 4 | - |
| $b$ | Birth rate | 342.5 day$^{-1}$ | - |
| $1/m$ | Maternal antibody duration | 2.9 months | [2] |
| $k$ | MCV dose | 1, 2 | - |
| $v_i^{(k)}$ | Age specific dose $k$ conditional probability | Equation 8 | - |
| $VE_i^{(k)}$ | Age specific dose $k$ VE | VE SCAM | [3] |
| $i_0^{(1)}$ | Recommended MCV1 age | 6–20 months | - |
| $i_0^{(2)}$ | Recommended MCV2 age | $i_0^{(2)} = i_0^{(1)} + 6$ | - |
| $\kappa_1$ | MCV1 vaccine coverage | 45%–85% | [4] |
| $\kappa_2$ | MCV2 vaccine coverage | $\kappa_2 = \kappa_1 - 5\%$ | [4] |
| $\lambda_i$ | Age specific force of infection | Equation 11 | - |
| $c_{i,j}$ | Contact rate between age groups $i$ and $j$ | Values from SCMs | [5, 6] |
| $q$ | <5 years transmissibility | 0.005–53.1 | Fitted |
| $R_0$ | Basic reproduction number | 10–20 | [7] |
| $1/\gamma$ | Infectious period | 13 days | [8] |

Table 1: Model parameters used in the measles transmission and vaccination model.

| Location | Age (years) | Time period | Transmission level |
|---|---|---|---|
| England and Wales | 4.5 – 5.5 | 1944 – 1960 | Low |
| Various localities in North America | 4.0 – 6.0 | 1912 – 1918 | Low |
| Zambia, Rhodesia, and South Africa | 3.0 – 4.0 | 1960 – 1968 | Medium |
| Nepal (Terai) | 3.0 – 4.0 | 1977 | Medium |
| Ghana | 2.0 – 3.0 | 1960 – 1968 | High |
| Eastern Nigeria | 2.0 – 3.0 | 1960 – 1968 | High |
| India (Pondicherry) | 2.0 – 3.0 | 1978 | High |
| Morocco | 2.0 – 3.0 | 1960 | High |

Table 2: Historical pre-vaccine mean ages of measles infection by location [9].

| Fixed effects | | | | | |
|---|---|---|---|---|---|
| | Estimate | Standard Error | 95% CI | t-value | F-value |
| Intercept | 12.6 | 0.84 | (10.8, 14.3) | 14.9 | |
| Low → medium | -1.6 | 0.02 | (-1.6, -1.5) | -77.8 | 15373 |
| Low → high | -3.6 | 0.02 | (-3.6, -3.5) | -175.0 | 15373 |
| **Random Effects** | | | | | |
| | Type | | Variance | Standard deviation | |
| SCM | Intercept | | 2.7 | 1.6 | |
| MCV1 coverage | Intercept | | 1.6 | 1.3 | |

Table 3: Coefficients of a linear mixed effects model of optimal age predicted by transmission level. Model equation: Optimal age $\sim$ Transmission level $+ (1|\,\text{SCM}) + (1|\,\text{MCV1 coverage})$. Confidence intervals (CI) calculated using likelihood ratio testing.

| Fixed effects | | | | | |
|---|---|---|---|---|---|
| | Estimate | Standard Error | 95% CI | t-value | F-value |
| Intercept | 3.4 | 1.2 | (0.8, 6.0) | 2.9 | |
| MCV1 coverage | 0.1 | 0.001 | (0.1, 0.2) | 118.1 | 13942 |

| Random Effects | | | |
|---|---|---|---|
| | Type | Variance | Standard deviation |
| SCM | Intercept | 2.6 | 1.6 |
| Transmission level | Intercept | 3.1 | 1.8 |

Table 4: Coefficients of a linear mixed effects model of optimal age predicted by MCV1 coverage. Model equation: Optimal age $\sim$ MCV1 coverage + (1| SCM) + (1| Transmission level).

### 3 Supplementary figures

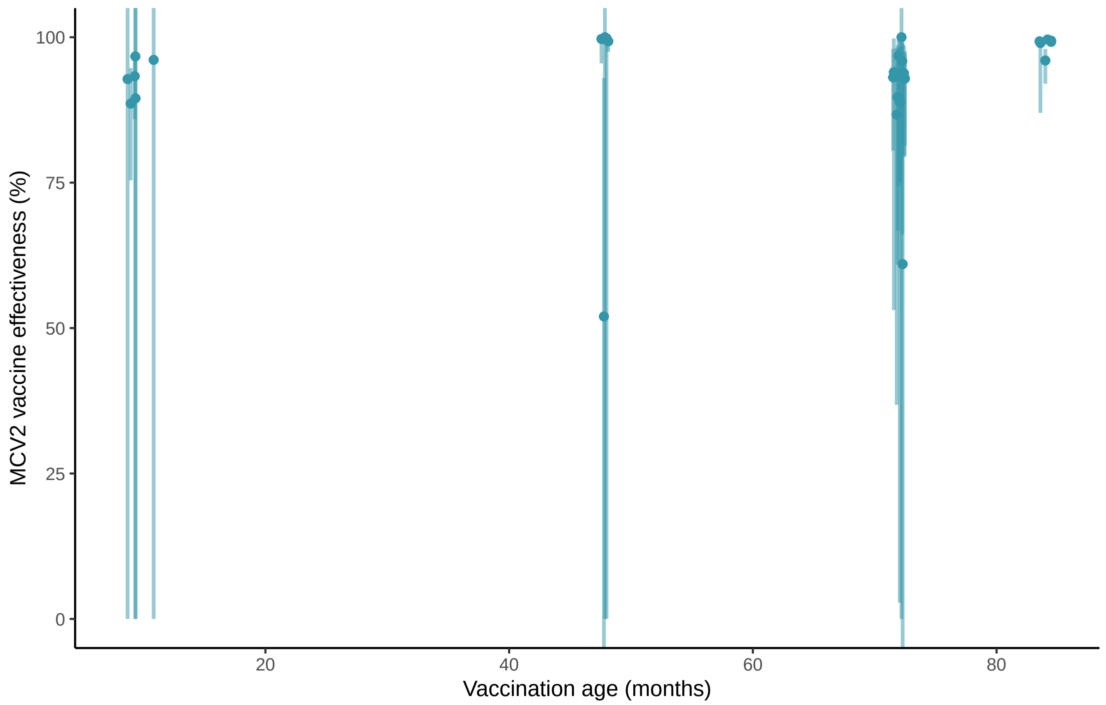

Figure 1: Estimates of vaccine effectiveness for Measles Containing Vaccine Dose 2, extracted from [3].

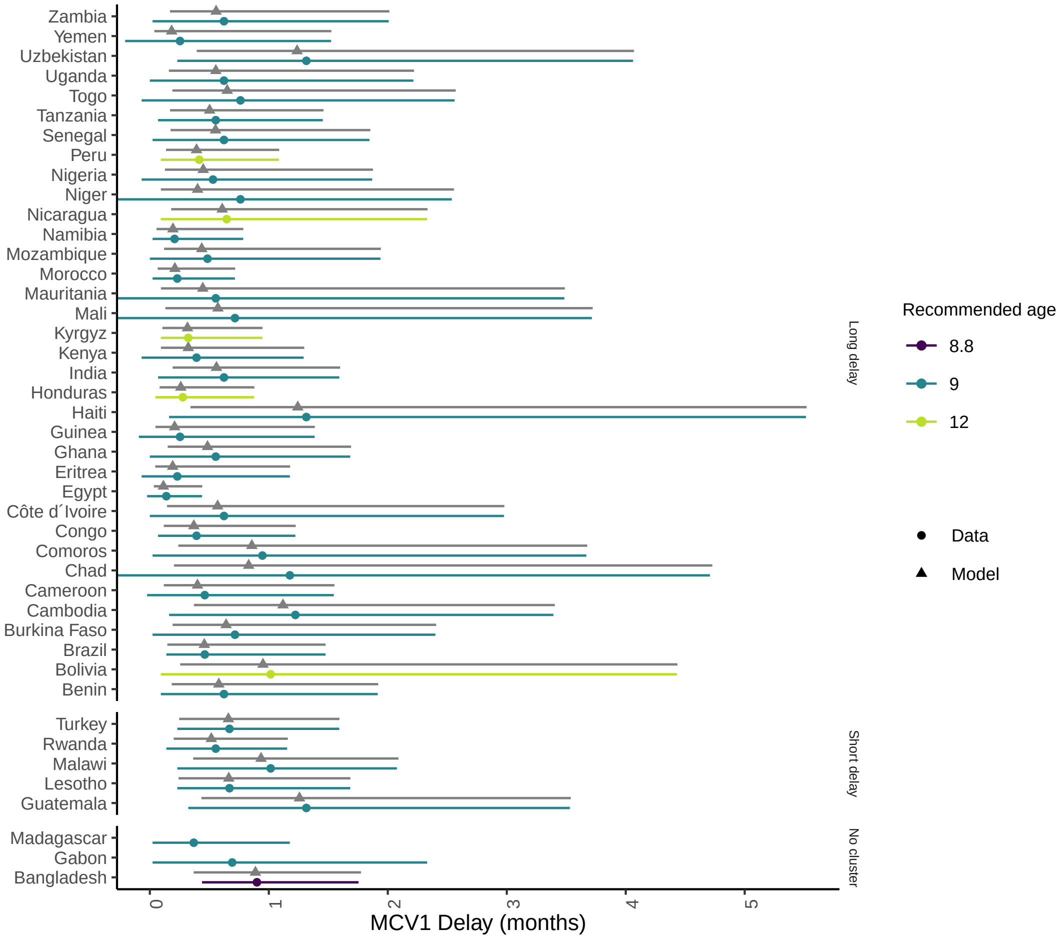

Figure 2: Measles Containing Vaccine Dose 1 delay data and modeled Lomax distribution, grouped by cluster. Points indicate median delay, and line segments indicate the 25% and 75% quantiles. Data are indicated by circles, modeled results indicated by triangles.

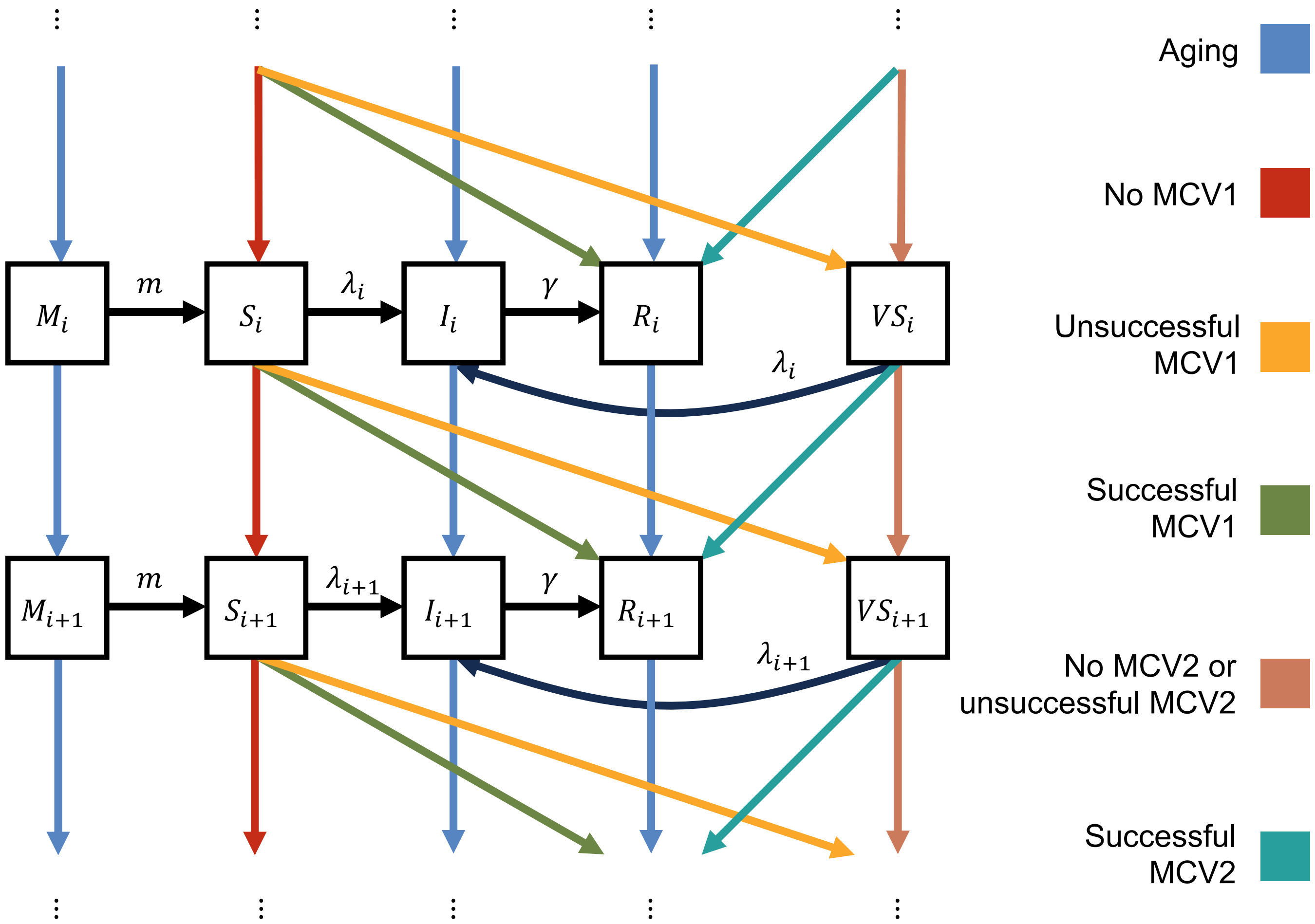

Figure 3: Model of measles transmission and vaccination schematic. Vaccination-related transitions are indicated by colours, and occur between age groups. Transmission-related transitions are indicated by black arrows, and occur within age groups.

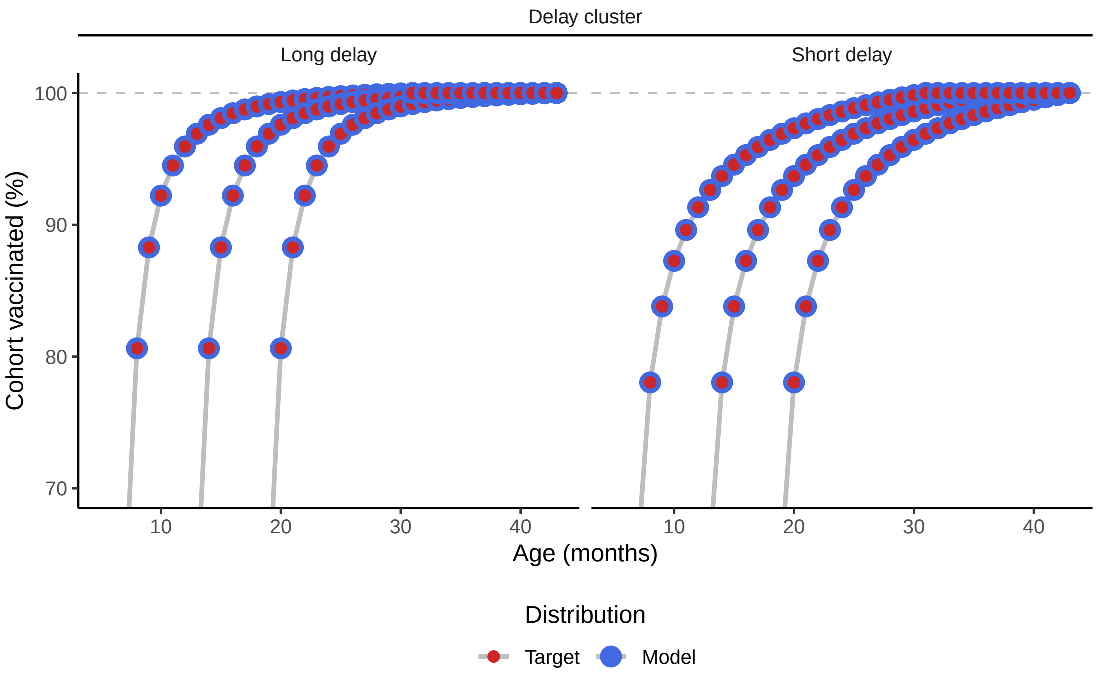

Figure 4: Comparison of the percentage of the population vaccinated with MCV1 by a given age according to the target distribution, derived from the delay distribution, and according to the measles transmission and vaccination model (detailed in 1.2.1).

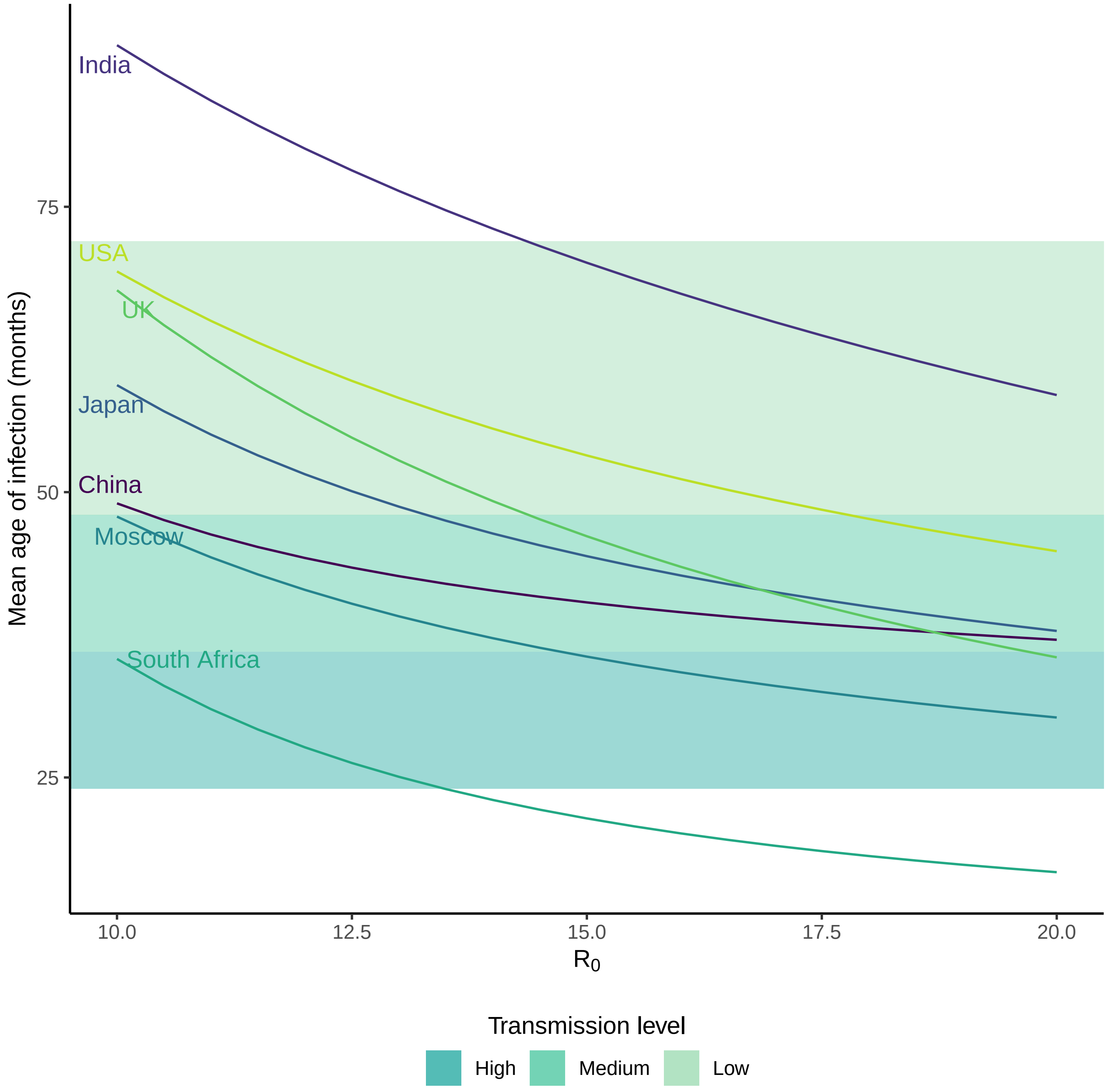

Figure 5: Modeled mean age of measles infection for different Social Contact Matrices (SCMs), at typical reported values of $R_0$ [7]. The mean age of infection values were calculated using the model with relative <5 years transmissibility, $q = 1$. Transmission levels were defined using historical reports of the mean age of infection [9].

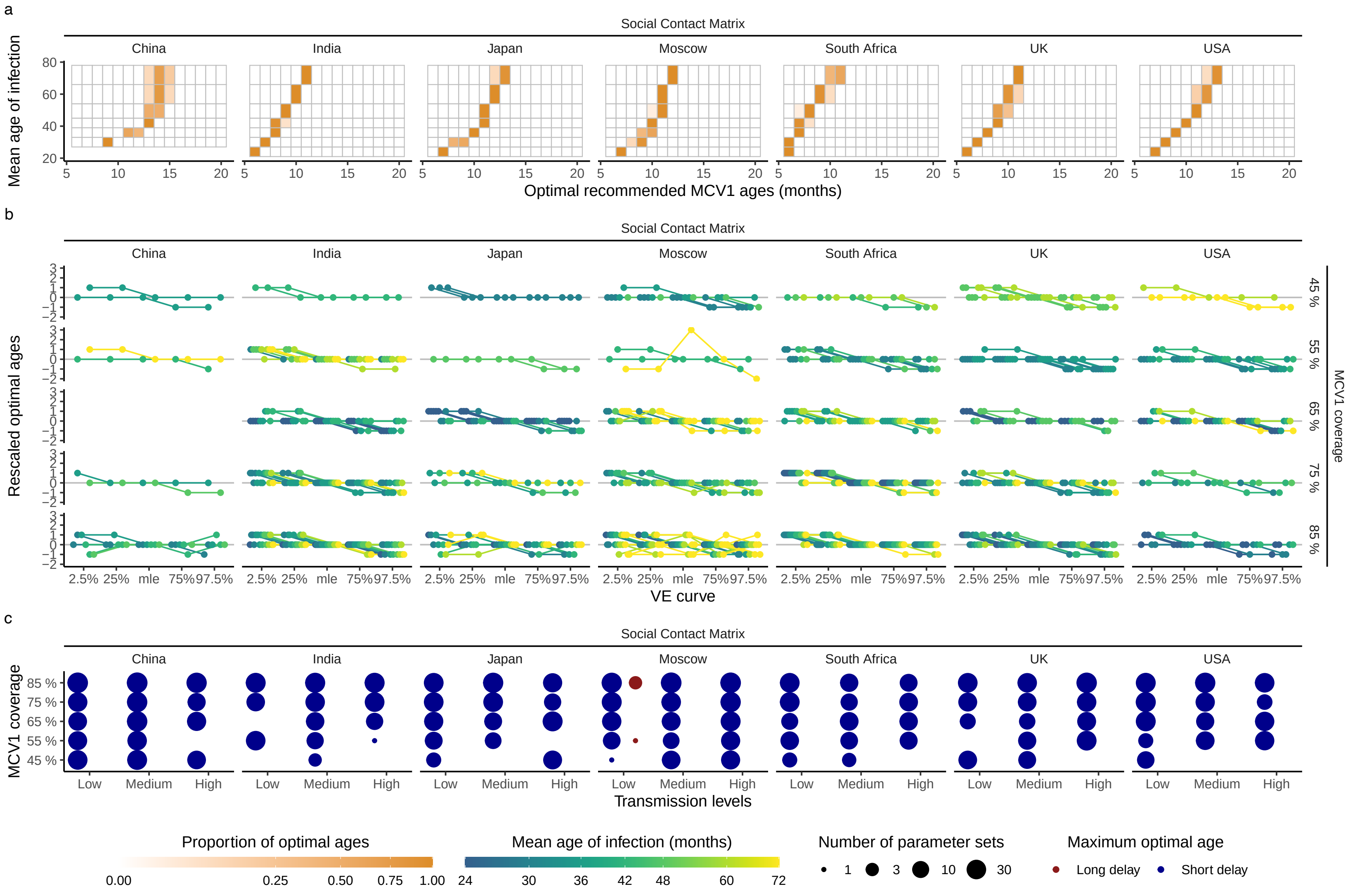

Figure 6: a) Impact of the mean age of infection on the optimal MCV1 age for each SCM. b) Impact of MCV1 VE curve on the optimal MCV1 age. Only parameter sets where changing the VE curve resulted in changes in optimal age are plotted. c) Impact of vaccination delay distribution on the optimal age. Only parameter sets with changes in optimal age are plotted.

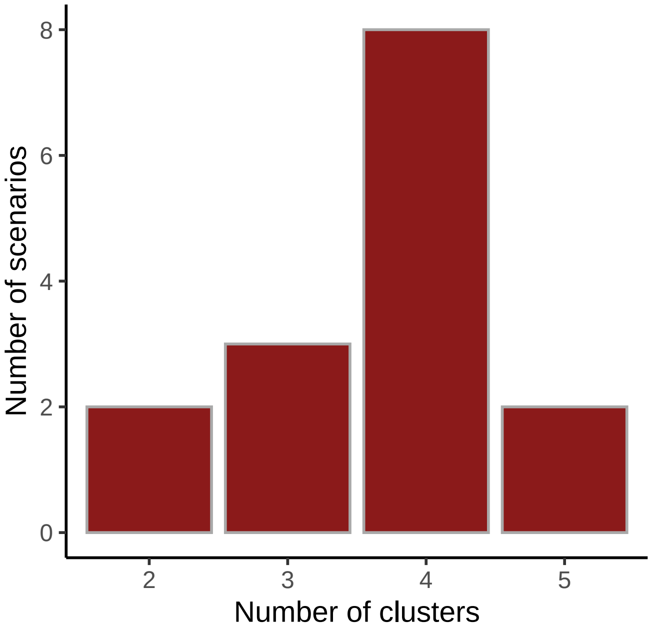

Figure 7: Histogram of the distribution of number of clusters. For each MCV coverage-transmission level pair (scenario) the optimal MCV1 ages were clustered according to Social Contact Matrix (SCM), using Partitioning Around Medoids (PAM) clustering with number of clusters determined using the silhouette method [10].

[4] World Health Organisation. WHO immunization data portal. https://immunizationdata.who.int.

[10] Leonard Kaufman and Peter J. Rousseeuw. *Finding Groups in Data.* Wiley Series in Probability and Statistics. John Wiley & Sons, Inc., New Jersey, 2005.
